# Black and Native Overdose Mortality Overtook that of White Individuals During the COVID-19 Pandemic

**DOI:** 10.1101/2021.11.02.21265668

**Authors:** Joseph Friedman, Helena Hansen

**Affiliations:** Medical Scientist Training Program, University of California, Los Angeles; Center for Social Medicine and Humanities, University of California, Los Angeles

## Abstract

Drug overdose mortality rates have increased sharply during the COVID-19 pandemic. In recent years, overdose death rates were rising most rapidly among racial/ethnic minority communities. The pandemic has disproportionately affected communities of color in a wide swath of health, social, and economic outcomes. Careful attention is therefore warranted to trends in overdose mortality by race/ethnicity during COVID-19. We calculated total drug overdose death rates per 100,000 population by race/ethnicity for the 1999-2020 time period. We find that Black overdose mortality overtook that of White individuals in 2020 for the first time since 1999. Between 2019 and 2020 Black individuals had the largest percent increase in overdose mortality, of 48.8%, compared to 26.3% among White individuals. In 2020, Black overdose death rates rose to 36.8 per 100,000, representing 16.3% higher than the rate for White individuals for the same period. American Indian and Alaska Native (AI/AN) individuals experienced the highest rate of overdose mortality in 2020, of 41.4 per 100,000, representing 30.8% higher than the rate among White individuals. Our findings suggest that drug overdose mortality is increasingly becoming a racial justice issue in the United States and appears to have been exacerbated by the COVID-19 pandemic. Providing individuals with a safer supply of drugs, closing gaps in access to MOUD and harm reductions services, and ending routine incarceration of individuals with substance use disorders represent urgently needed, evidence-based strategies that can be employed to reduce rising inequalities in overdose.

## Introduction

Drug overdose mortality rates have increased sharply during the COVID-19 pandemic^1^. In recent years, overdose death rates were rising most rapidly among racial/ethnic minority communities^2^. The pandemic has disproportionately affected communities of color in a wide swath of health, social, and economic outcomes. Careful attention is therefore warranted to trends in overdose mortality by race/ethnicity during COVID-19.

## Methods

We calculated total drug overdose death rates per 100,000 population by race/ethnicity for the 1999-2020 time period, using data obtained from the National Center for Health Statistics^3^. Records from 2020 were provisional and may underestimate the final level of drug overdose related mortality. Drug overdose deaths were classified as those assigned with the underlying cause of death in the ICD-10 categories pertaining to unintentional, suicide, homicide, or undetermined intent drug overdose deaths, (X40-44, X60-64, X85, or Y10-14, respectively). We calculated annual percent change in a race/ethnicity specific fashion for the 2000-2020 period. All analyses were conducted using R version 4.0.3. See appendix for more details.

## Results

We find that Black overdose mortality overtook that of White individuals in 2020 for the first time since 1999. In 2020, Black overdose death rates rose to 36.8 per 100,000, representing 16.3% higher than the rate for White individuals for the same period. This is a sharp reversal of the Black-White overdose mortality gap noted in 2010, when the rate among White individuals was double (100.1% higher) than that seen among Black individuals. These shifts reflect that Black communities have experienced higher annual percent increases in overdose deaths compared to their White counterparts each year since 2012. In 2020 Black individuals had the largest percent increase in overdose mortality, of 48.8%, compared to 26.3% among White individuals.

American Indian and Alaska Native (AI/AN) individuals experienced the highest rate of overdose mortality in 2020, of 41.4 per 100,000, representing 30.8% higher than the rate among White individuals. Between 1999 and 2017 AI/AN overdose mortality rates were close to those experienced by White individuals, with rates among AI/AN first overtaking those of Whites by a large margin in 2019. In 2020 overdose mortality increased by 43.3% for AI/AN individuals.

Drug overdose rates among Latinx individuals remained the lowest among the 4 race/ethnicity groups assessed throughout the study period, however they also experienced a large percent increase in 2020 of 40.1%.

For all race/ethnicity groups assessed, the relative increases observed in 2020 were higher than any prior percent change observed between 1999-2019.

## Discussion

The overdose crisis in the United States is increasingly driven by a toxic illicit drug supply characterized by polysubstance use of potent synthetic opioids and benzodiazepines, as well as high-purity methamphetamine. The high—and unpredictably variable—potency of the illicit drug supply may be disproportionately harming communities of color for various reasons. Deep-seated inequalities in living conditions including stable housing and employment, policing and arrests, preventive care, harm reduction, telehealth, medications for opioid use disorder (MOUD) and naloxone, are likely playing a key role^4,5^.

Further, the increasing toxicity of the drug supply has increased the lethality of recent incarceration—which disproportionately affects Black and Native individuals due to structural racism in the criminal justice system—as a risk factor for overdose mortality. Recently incarcerated individuals have reduced opioid tolerance and less knowledge of shifts in drug potency^6^.

Drug overdose mortality is increasingly becoming a racial justice issue in the United States and appears to have been exacerbated by the COVID-19 pandemic. Providing individuals with a safer supply of drugs, closing gaps in access to MOUD, health care, and harm reduction services, and ending routine incarceration of individuals with substance use disorders represent urgently needed, evidence-based strategies that can be employed to reduce rising inequalities in overdose.

**Figure 1.**
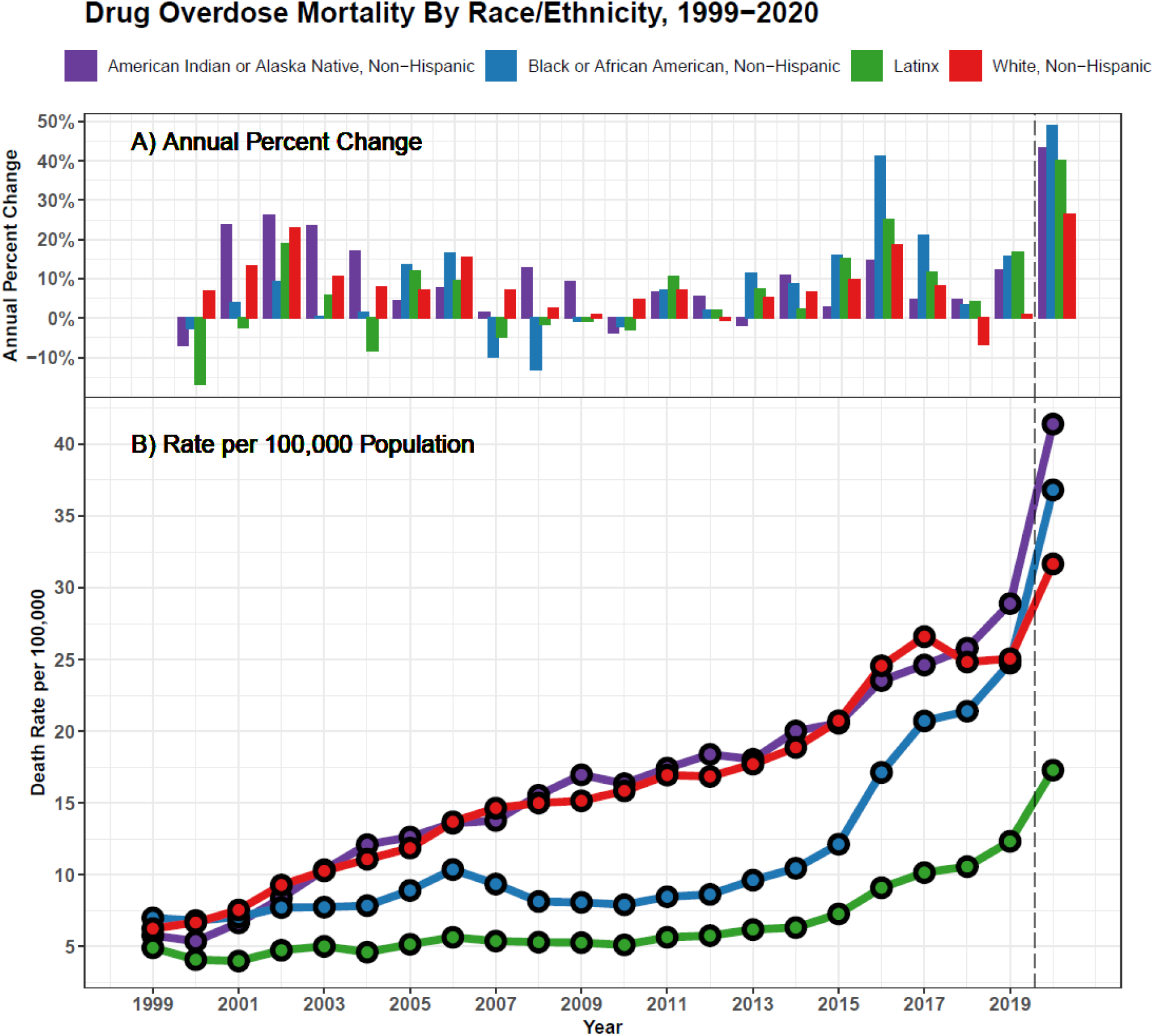
Drug Overdose Mortality by Race/Ethnicity, 1999-2020. Year-to-year percent change in drug overdose mortality by race/ethnicity (top). Drug overdose mortality per 100,000 population by race/ethnicity (bottom). A vertical dashed line separates the COVID-19 pandemic period from prior trends.

## Supporting information

Data Produced in Analysis

## Data Availability

All data produced in the present work are contained in the supplemental files released with the manuscript.

## Methods Appendix

### Data Preparation and Definitions

1. Total drug overdose mortality counts for the 1999-2019 period were obtained from the CDC WONDER platform (https://wonder.cdc.gov/controller/saved/D76/D243F427) stratified by race, ethnicity, and year of occurrence. Both population counts, and counts of overdose fatalities were provided, and used as the denominator and numerator, respectively.
2. Overdose mortality rates for 2020 were calculated using provisional race/ethnicity stratified drug overdose mortality counts released by the National Center for Health Statistics (https://www.cdc.gov/nchs/data/health_policy/Provisional-Drug-Overdose-Deaths-by-Quarter-and-Demographic-Characteristics-2019-to-2020.pdf).
3. Mid-year population estimates by race/ethnicity for the year 2020 were obtained from the CDC Wonder platform (https://wonder.cdc.gov/single-race-single-year-v2020.html).
4. Total drug overdoses were defined by the following ICD-10 codes: 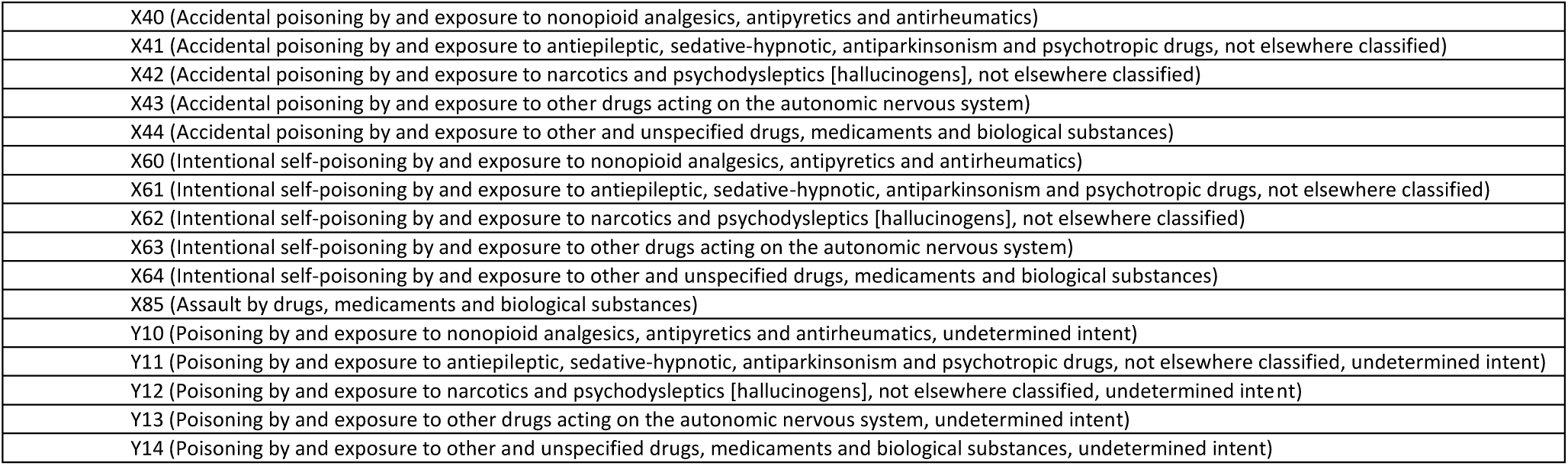
5. ‘Latinx’ individuals were defined as any persons for which ethnicity was defined as Hispanic or Latino, regardless of race. “American Indian or Alaska Native, Non-Hispanic,” “Black or African American, Non-Hispanic,” and “White, Non-Hispanic” individuals were defined as individuals of each race, who had their ethnicity listed as “non-Hispanic.”
6. Rates were calculated as total drug overdose deaths per 100,000 population, according to mid-year population.
7. Year-to-year percent change values were calculated for each year *t* in a race/ethnicity group-specific fashion according to the following formula: *((Overdose Death Rate_*t*_ /Overdose Death Rate*_*t-1*_*)-1)*100%*.
8. Data were visualized using R version 4.0.3.

### Methodological Consideration

1. Records from 2020 were provisional and may underestimate the final level of drug overdose related mortality.
2. Race/ethnicity may be incorrectly assigned in some overdose deaths. This is a known issue especially for Native individuals.
3. It was not possible to age-standardize the race/ethnicity stratified results given the format of the provisional records from 2020. This is an important area of consideration for future work.
4. This work is descriptive, not causal in nature, and it was not possible to show causally what percent of increases in overdose death rates between 2019 and 2020 stem from the COVID-19 pandemic directly.

## References

1. Friedman J, Akre S. COVID-19 and the Drug Overdose Crisis: Uncovering the Deadliest Months in the United States, January-July 2020. Am J Public Health. Published online April 15, 2021:e1–e8. doi:10.2105/AJPH.2021.306256

2. James K, Jordan A. The Opioid Crisis in Black Communities. J Law Med Ethics. 2018;46(2):404–421. doi:10.1177/1073110518782949

3. National Center for Health Statistics. Provisional Drug Overdose Deaths by Quarter and Demographics - 2019 to 2020. Published online 2021. Accessed October 23, 2021. https://www.cdc.gov/nchs/data/health_policy/Provisional-Drug-Overdose-Deaths-by-Quarter-and-Demographic-Characteristics-2019-to-2020.pdf

4. Lagisetty PA, Ross R, Bohnert A, Clay M, Maust DT. Buprenorphine Treatment Divide by Race/Ethnicity and Payment. JAMA Psychiatry. 2019;76(9):979–981. doi:10.1001/jamapsychiatry.2019.0876

5. Park JN, Rouhani S, Beletsky L, Vincent L, Saloner B, Sherman SG. Situating the Continuum of Overdose Risk in the Social Determinants of Health: A New Conceptual Framework. The Milbank Quarterly. 2020;98(3):700–746. doi:10.1111/1468-0009.12470

6. Brinkley-Rubinstein L, Macmadu A, Marshall BDL, et al. Risk of fentanyl-involved overdose among those with past year incarceration: Findings from a recent outbreak in 2014 and 2015. Drug and Alcohol Dependence. 2018;185:189–191. doi:10.1016/j.drugalcdep.2017.12.014

